# Quantifying Covid19-Vaccine Location Strategies For Germany

**DOI:** 10.1101/2020.11.18.20234146

**Authors:** Neele Leithäuser, Johanna Schneider, Sebastian Johann, Sven O. Krumke, Eva Schmidt, Manuel Streicher, Stefan Scholz

## Abstract

**Background:** Vaccines are an important tool to limit the health and economic damage of the Covid-19 pandemic. Several vaccine candidate already provided promising effectiveness data, but it is crucial for an effective vaccination campaign that people are willing and able to get vaccinated as soon as possible. Taking Germany as an example, we provide insights of using a mathematical approach for the planning and location of vaccination sites to optimally administer vaccines against Covid-19.

**Methods:** We used mathematical programming for computing an optimal selection of vaccination sites out of a given set (i.e., university hospitals, health department related locations and general practices). Different patient-to-facility assignments and doctor-to-facility assignments and different constraints on the number of vaccinees per site or maximum travel time are used.

**Results:** In order to minimize the barriers for people to get vaccinated, i.e., limit the one-way travel journey (airline distance) by around 35 km for 75 % of the population (with a maximum of 70 km), around 80 well-positioned facilities can be enough. If only the 38 university hospitals are being used, the 75 % distance increases to around 50 km (with a maximum of 145 km). Using all 400 health departments or all 56 000 general practices can decrease the journey length significantly, but comes at the price of more required staff and possibly wastage of only partially used vaccine containers.

**Conclusions:** In the case of free assignments, the number of required physicians can in most scenarios be limited to 2 000, which is also the minimum with our assumptions. However, when travel distances for the patients are to be minimized, capacities of the facilities must be respected, or administrative assignments are prespecified, an increased number of physicians is unavoidable.

## Introduction

In the expectation of an upcoming availability of vaccines against the SARS-CoV-2 virus, public health authorities already need to make appropriate preparations in order to utilize the available vaccine capacities from the very beginning. In politics, there are already numerous guidelines available on how to deal with such a large-scale vaccination which affects all countries simultaneously [2, 7, 6, 25, 24]. Besides from analyzing the actual number of people willing to get a vaccination [14, 10], one crucial decision, that researchers around the world currently study is the question of who to vaccinate first in the face of scarce resources (cf. [15, 3]). Our focus however lies on the logistic decision on where to administer the vaccine to the people. Aside from operational aspects, such as cooling the vaccination doses, availability of medical staff, or spacious waiting areas that allow social distancing, the people’s journey duration is an important factor to take into account, as it is assumed that it correlates with the willingness to be vaccinated [4, 7, 16].

The most convenient way of vaccine administration for the population would be a vaccination at the local physician as it is common for other vaccinations such as influenza or tetanus. However, given the expected shortness of doses in comparison to the demand in the initial phase of the vaccination campaign, such a decentralized setting on the one hand poses the risk of wastage due to fixed container sizes and non-adherence to vaccination recommendations on the other hand. The biggest disadvantages, however, are the technical aspects: In the most likely scenario of starting with mRNA-based vaccinations, the vaccine must be transported at about −70 °C and stored at −20 °C [5, 20], which may require cooling equipment or special logistics that cannot be provided by most physician practices.

In the absence of knowledge of the actual temperature and other technical requirements, the well-equipped university hospitals were therefore brought into discussion as possible vaccination sites. Since there are only 38 of them in Germany, it is obvious that the distances for the vast majority of citizens are significantly longer than in the general physician scenario. Bundling those to be vaccinated in a central location makes it easier to distribute the vaccinations only to the intended population cohorts; at the same time, however, many more people, especially from risk groups, meet in one place. In times of a high incidence, this increases the probability of a disease transmission during at the vaccination site running contrary to the intention of vaccination.

A compromise scenario is vaccinating the people in public health departments, which are also decentralised and could be equipped at least with less specialised cooling technology. Since on average several hundred people have to be vaccinated every day, it is still necessary to create a safe hygiene concept. Most likely, the vaccinations itself will not be performed in the health department building, but rather, e.g., in gymnasiums or other adjoining event halls, since the health departments themselves usually do not have the necessary space for waiting areas or the like. This interpretation is also valid in our greenfield studies, where arbitrary facilities in the area could be used.

This study aims to find long term plans for locations of vaccination centers and assignments of patients to these centers within a given fixed time frame. We do not aim to give detailed information on which patient is vaccinated at which center at what exact moment in time. Rather, we want to give information on how many patients from a fixed given area, such as municipalities, are to be vaccinated at which center in a fixed time frame, such as a week.

The models and methods used in this study can be adjusted to fit any given area and sets of possible vaccination centers. It would therefore be quite possible to provide detailed solutions for, say, a certain state within Germany, provided data about possible vaccination centers is provided. The possible centers chosen for the study below demonstrate the applicability of our models and solution approaches to the problem of distributing vaccinations.

This study aims at quantifying the key indicators *required staff* and *journey lengths* in different scenarios that are currently discussed by German decision makers. These are (I) vaccinating decentralized at local general practices, vaccinating at public health departments throughout the country, or (III) vaccinating at few, but highly equipped university hospitals. Based on the health department locations of (II), which more or less correspond to counties, we also consider (IV) a greenfield approach, where we open only a subset of locations.

This paper is structured as follows: First we give the details of the study setup, its parameters and considered objectives. Then, we give the results for the four scenarios (I) to (IV) followed by a short discussion on the sensitivities of our parameters and methods. In order to make it more readable for non-mathematicians, we defer the mathematical background to the end, where we explain the used methodology and give insights into the strength of the used models. We close the paper with an outlook on further aspects that yet have to be integrated into the model.

## Methods

In order to get a sensible trade-off between complexity and significance, we decided upon using all German municipalities as basis regions for our evaluations and consider only linear distances between the municipality’s reference coordinate and a vaccination facility. The main variable in our model is the decision of which proportion of the municipality’s population should be sent to which vaccination facility. Depending on the specific scenario this variable often depends on the linear distance between the municipality and the vaccination facility, but can also be fixed beforehand by a given assignment. For each municipality, we calculate the part of its population to be vaccinated in the given time frame by scaling the total number of vaccination doses by the municipality’s population size. Here, it is also possible to consider, e.g., only elderly people or system-critical employees. In this way, we ensure that the number of vaccinated citizens is distributed fairly across all regions. We also assume that each physician has a maximum number of vaccinations that he can perform in a week. In some scenarios we consider lower and upper bounds for the total number of patients that can be assigned to one vaccination center. These bounds can be used to avoid overloading a specific vaccination center as well as to ensure a fair distribution of the patients to the centers.

Beyond the above setting, the model is very versatile. In particular, it can handle a lot of different scenarios, i.e., we may choose different types of regions, vaccination centers, assignments between the regions and vaccination centers, etc. All in all, we formulate three questions for each scenario:

1. Which vaccination centers should be opened?
2. How many physicians are needed in each vaccination center?
3. Which citizens should be vaccinated in which vaccination center?

We always aim to answer these questions with specific goals in mind. Examples for these objectives are:

- Minimizing the number of open stations,
- minimizing the sum of the distances the patients have to travel for vaccination, or
- minimizing the number of physicians needed.

Here it is important that we can pursue not only one objective, but any lexicographic combination of these objectives. Other possible objectives may easily be added into the model. Note that we model physicians as integral variables that is, we count heads not full-time equivalents. Otherwise, the total (continuous) number of physicians would be constant for all assignments. Minimizing the number of physicians per facility is not only important for logistic reasons (a physician may not be able to travel amongst different locations) and overhead costs (such as instruction courses, personal equipment, etc.), but can also directly be linked to the number of vaccine containers that will be opened and should not be wasted. In the presented studies, we assume that a doctor’s weekly vaccination capacity is a multiple of the total number of doses in a container.

To compute answers to the raised questions with the help of our model we need some assumptions and data. In our studies we consider a weekly cycle. Our personal correspondence with Stefan Scholz from the RKI has shown that their current estimate is to have around 500 000 vaccine doses available per week. They further assume that a doctor needs about 10 minutes on average for a vaccination and can therefore vaccinate 250 patients in a 5-day week of 8 hours each. The theoretical minimum is therefore a total of 500 000*/*250 = 2 000 physicians. Further, we received data about university hospitals and health departments from the RKI. We have obtained the population data of the municipalities in Germany from the federal statistical office (Statistisches Bundesamt) in Germany, cf. [21]. The data concerning general practices has been collected by Stefan Scholz and Katharina Schmidt in 2013 using publicly available data and can be found on the web, cf. [26]. Further, data on the age structures in Germany is provided on a website of the Bundesamt für Bauwesen und Raumordnung, cf. [11]. Since our main concern is to compare the resulting proportions of the key figures, the different ages of the data are not problematic.

We also consider the whole German population as the reference population that should be vaccinated. In Section Discussion, we discuss the sensitivity of the results to the weekly vaccination capacity, the weekly doses of vaccine available and partial populations.

According to the current political discussions and our personal communication with the RKI, the doses shall be distributed among the states according to their population size as shown in Table 1 [2].

**Table 1:** Distribution of weekly vaccinations per state.

|  | State | Abbreviation | Population | Vaccinations |
| --- | --- | --- | --- | --- |
| 1 | Baden-Wuerttemberg | BW | 11 069 533 | 66 670 |
| 2 | Bavaria | BY | 13 076 721 | 78 761 |
| 3 | Berlin* | BE | 3 644 826 | 21 952 |
| 4 | Brandenburg | BB | 2 511 917 | 15 134 |
| 5 | Bremen* | HB | 682 986 | 4 113 |
| 6 | Hamburg* | HH | 1 841 179 | 11 089 |
| 7 | Hesse | HE | 6 265 809 | 37 734 |
| 8 | Mecklenburg-Vorpommern | MV | 1 609 675 | 9 697 |
| 9 | Lower Saxony | NI | 7 982 448 | 48 071 |
| 10 | North Rhine - Westphalia | NW | 17 932 651 | 108 005 |
| 11 | Rhineland Palatinate | RP | 4 084 844 | 24 591 |
| 12 | Saarland | SL | 990 509 | 5 965 |
| 13 | Saxony | SN | 4 077 937 | 24 569 |
| 14 | Saxony-Anhalt | ST | 2 208 321 | 13 297 |
| 15 | Schleswig-Holstein | SH | 2 895 229 | 17 440 |
| 16 | Thuringia | TH | 2 143 145 | 12 912 |
|  | **Total** |  | **83 017 730** | **500 000** |

\*city states with BE and HH consisting of only one and HB of two municipalities.

### Vaccination in General Practices (I)

The general medical care is one of the cores of the German health care system. As a rule, every German citizen has compulsory health insurance and pays a health insurance contribution, which depends on a percentage of the income from employment. In return, everyone is entitled to free medical services at any time and can consult any doctor who is authorized to treat them under the statutory health insurance system. In our model we assume that there are about 56 000 general practitioners in Germany, where this data is from 2013, see [26]. Of course, this number has changed in the meantime, but this is where the advantages of our model come into play. Namely if we obtain newer data sets, we can easily add them and adjust our results. The general practices are distributed in such a way that one can usually reach the nearest one within around 5 km, cf. [13]. In addition to the easy accessibility of the general practitioners, they also carry out general vaccinations. Therefore, general practices are a suitable and reasonable choice for vaccination centers.

As mentioned earlier, if not stated otherwise, we always assume that there are 500 000 available vaccines each week and that one general practitioner can carry out 250 vaccinations during this time. We model multiple licensed doctors within a joint practice or within the same building as separate facilities with separate capacities.

In the case of general practitioners providing the vaccinations we consider the following scenarios:

> ***free_assignment*** In this scenario, we are interested in which the general practitioners should carry out vaccinations and where patients are vaccinated. The first objective of these calculations is to open as few vaccination centers as possible, allowing a maximal distance of 50 km per patient. After optimizing this first objective, we also minimize the total sum of distances the patients have to travel to get their vaccination.

> ***free_assignment_only_distance*:** Each patient is assigned to his closest practice possible such that the physician’s capacity is not exceeded. Therefore, we minimize the sum of the travel distances of all patients while also respecting that each doctor can carry out at most 250 vaccines. As a secondary objective, we minimize the number of open practices, in order to reduce underutilized physicians that would lead to vaccine wastage.

Note that the scenario of each person choosing her closest physician would lead to overcrowded practices at some places, where more than 250 patients would choose the same doctor. However, the resulting extra travel distance in comparison to *free_assignment_only_distance* is negligible.

If vaccines need to be stored in dry ice to keep the cooling requirements, it is only reasonable to perform vaccines for a multiple of the package size. If the size is e.g. 250, we need exactly to fill up the physician’s capacity in order to utilize all shots. The vaccine spoils after a short number of days after opening the package. Since the vaccine is expected to be very scarce at the beginning, wastage is a very undesirable outcome. By bundling doctors and limiting the number of practices, the effect can immanently be limited. Opening more or all facilities than necessary without further time reduction would only lead to more wastage.

### Vaccination at Public Health Departments (II)

Health departments are the local authorities that are part of the German public health service. They are responsible for the execution of the medical tasks of the health administration. In Germany there are almost 400 health departments and their duties are defined by federal laws, federal state laws and federal state regulations. Each municipality is usually assigned to exactly one health department, which in turn is assigned to exactly one federal state. Exceptions are the city states Berlin and Hamburg, which are modeled as big municipalities due to the used input data. However, due to their population size, they have in fact several health departments and it would thus be reasonable to subdivide them into city districts. Vice versa, most of the authorities are responsible for exactly one county. Some health departments are responsible for two counties, usually a city county and its surrounding county.

If a municipality is in a certain federal state, then it is assigned to a health department in the same federal state. The federal states are partly autonomous and hence take over the tasks of the health services for their citizens.

Health departments are already responsible for many health-related tasks and are therefore an obvious choice for vaccination centers. In the following scenarios we again assume that there are 500 000 vaccinations weekly available and each physician can vaccinate 250 people in a week:

> ***responsible_station:***The federal structure suggests that each resident of a municipality is vaccinated at the responsible health department. By design, the responsible station is always in the patient’s federal state. The open vaccination centers as well as the assignment from patients to vaccination center here is already given. Thus, we are interested in the number of physicians needed in every health department to be able to carry out all the vaccinations.

> ***closest_station_greedy:*** Sometimes the nearest public health department is not the one responsible for the municipality. For comparison we consider a scenario that neglects these described administrative structures. Here each citizen visits the closest health department whether it is in the same federal state or not. Once again the open vaccination centers and the assignment from citizens to vaccination centers are predefined. It remains to determine how many physicians are needed at each health department.

Due to a lack of data, we do not consider capacities of the individual health departments in terms of patients or doctors yet. However, if provided, this is easily integrable into our model.

### Vaccination at University Hospitals (III)

Another possibility that can be considered for vaccination centers are hospitals. Here we consider university hospitals in particular as they are the largest and most modernly equipped hospitals. There are 38 university hospitals spread all over Germany which are mainly located in large cities. The connection between rural areas and the big cities is mostly well maintained, but in some cases there are municipalities more than 100 km away from the closest university hospital.

However, in the following, we consider the university hospitals as possible vaccination centers. As before, we assume that 500 000 vaccines are available each week and that one physician can carry out 250 vaccinations per week. With these assumptions we consider three scenarios:

> ***closest_station_greedy*:** First we assume that vaccinations are carried out at all university hospitals and that every citizen is vaccinated at the nearest clinic. Further, we assume for this scenario that there are no capacities on the number of vaccinations that can be carried out in each university hospital. This can again be interpreted as the free choice of doctor. A solution to this scenario gives a rough overview on the distribution of the vaccine and the number of physicians needed at each university hospital.

> ***closest_station_same_state:*** This scenario is very similar to the previous one, but this time the federal structures of Germany are taken into account. In particular, every citizen has to visit a university clinic located in the same federal state he/she lives in. However, if there is no university hospital in a state, we allow the affected citizens to be vaccinated in a different state. Again we compute the number of physicians needed at every university hospital.
>
> ***free_assignment_only_distance:*** In contrast to the previous two scenarios, now we predefine upper and lower bounds of available vaccinations per clinic proportional to the university hospital’s size, or more precisely, to the number of outpatients they treat. We set the bounds to *±* 20% of this value. Thus, if the hospital opens as a vaccination center, the number of assigned citizens has to respect these bounds. In this case there are no valid solutions if every citizen goes to the nearest hospital. Therefore, we determine which citizen should be vaccinated at which university hospital with the objective to minimize the overall sum of travel lengths of the citizens.

### Greenfield Planning (IV)

At last, we study a free planning approach, where the number of facilities is dependent on a defined maximal acceptable radius.

For the sake of simplicity we use the coordinates of the health departments as potential set of vaccination facilities. We also assume that their geographical distribution roughly reflects Germany’s population density. We interpret those as arbitrary facilities and in particular, we do not open all potential facilities for vaccination. Observe that there are some (rural) municipalities which are not within a 50 km radius of a potential facility. These municipalities are assigned to their closest vaccination facility which therefore has to be open.

We study the following assignment variants with the same capacity assumptions as before:

***free_assignment_15km*** Each patient should be assigned to a location at a maximum distance of 15 km. Our primary goal is to minimize the number of open locations within that constraint. The secondary goal is to minimize the required number of physicians and at last, the actual travel distances are being minimized.

***free_assignment_30km*:** The setting is the same as above, but the bound is set to 30 km.

***free_assignment_50km*:** The setting is the same as above, but the bound is set to 50 km.

***free_assignment_75km*:** The setting is the same as above, but the bound is set to 75 km.

We also analyze the health department and the university hospital solutions on the level of federal states. This is reasonable, since the vaccine doses will probably be distributed among the states according to their population sizes. For the respective state governments, it is an undesired effect that their assigned vaccinations will be served to people from adjacent states. On the other hand, the convenience for their respective citizens will also be of concern. A comparison of results with or without crossing state borders may therefore lead to discussions about intergovernmental agreements, especially in the university hospital scenario, where there are states without any such facility.

## Results

In the following, we analyze each scenario according to the linear distance between the municipality centers and the assigned vaccination stations, the number of physicians and their capacity.

For various analyses throughout this section, we use boxplots as a mean of visualizing distribution information. We orient ourselves by the default setting [19] of refined boxplots, i.e. drawing the boxes as the lower and upper quantile around the median and drawing whiskers that specify 1.5 times the IQR (interquartile range) [22]. Data outside this range is identified as outliers and drawn as small diamonds.

First we look at vaccinations at general practices.

### Vaccination in General Practices (I)

The most convenient solution for the patients would of course be the vaccination at their general physician. But due to logistic requirements (cooling of the vaccine) and the prioritisation of population groups (e.g., people with high risk or with work-related high contact rates) this solution does not seems to be practicable in the beginning of the vaccination procedure. However, we still consider this scenario for the sake of comparison.

As described in Methods, we consider the following assignments for general practices:

i. ***free_assignment***
ii. ***free_assignment_only_distance***.

Due to the high density of physicians in Germany, mapping all locations would not be of any help. The map in Figure 1 however shows a reduced subset of the resulting open practices for the above assignment variants. For the sake of consistency with other maps later in this paper, the municipalities are also colored according to their median distance with respect to their assigned physicians. As we are limiting our analysis to the linear distances between the facilities location and the center point of a municipality, the detailed analysis of the distances is not very precise. It can however, easily be acknowledged that the vast majority of patients will be assigned to a rather close facility in both scenarios. By design, the distances are even smaller in assignment (ii), where this is our primary goal, although the presented solution is not proven to be optimal, since the running time exceeded the preset time constraints. The distribution of distances are visualized in Figure 2b. The main result in this scenario is that around 7 000 open practices out of 56 000 possible ones are enough to reach the best possible distance for the total population. Another important hint to deciders is that opening more than 2 000 locations automatically leads to a guaranteed wastage of vaccine doses if the number of shots per package is calibrated to the vaccination capacity of one full-time physician. This effect is visualized in Figure 2a.

**Figure 1:**
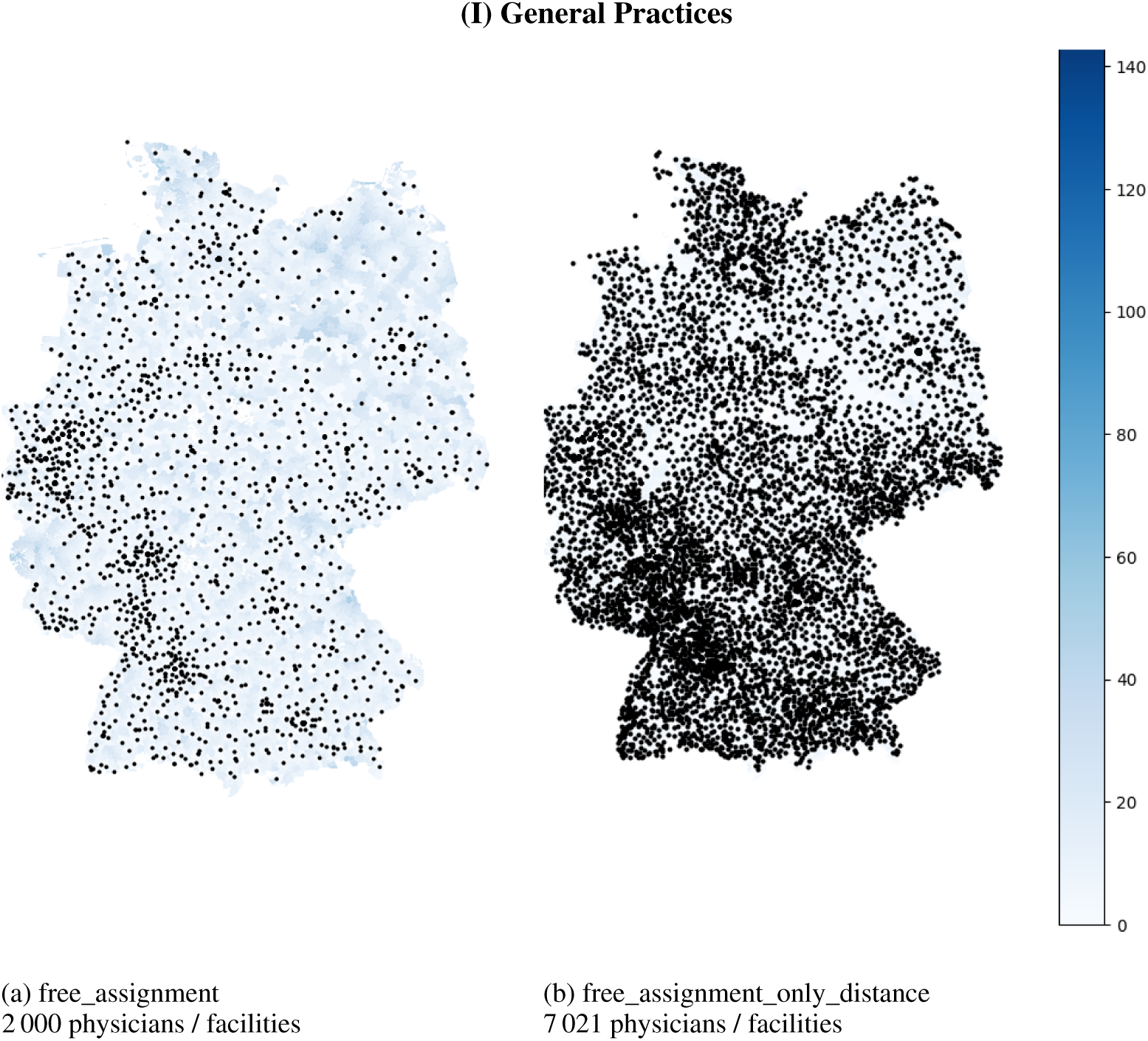
Visualization of the position of opened practices. The municipalities are colored with respect to their airline distance in kilometers to their assigned physician. As shown in Figure 2b, almost all municipalities are assigned to a practice closer than 10 km in the right figure and would be colored very lightly.

**Figure 2.**
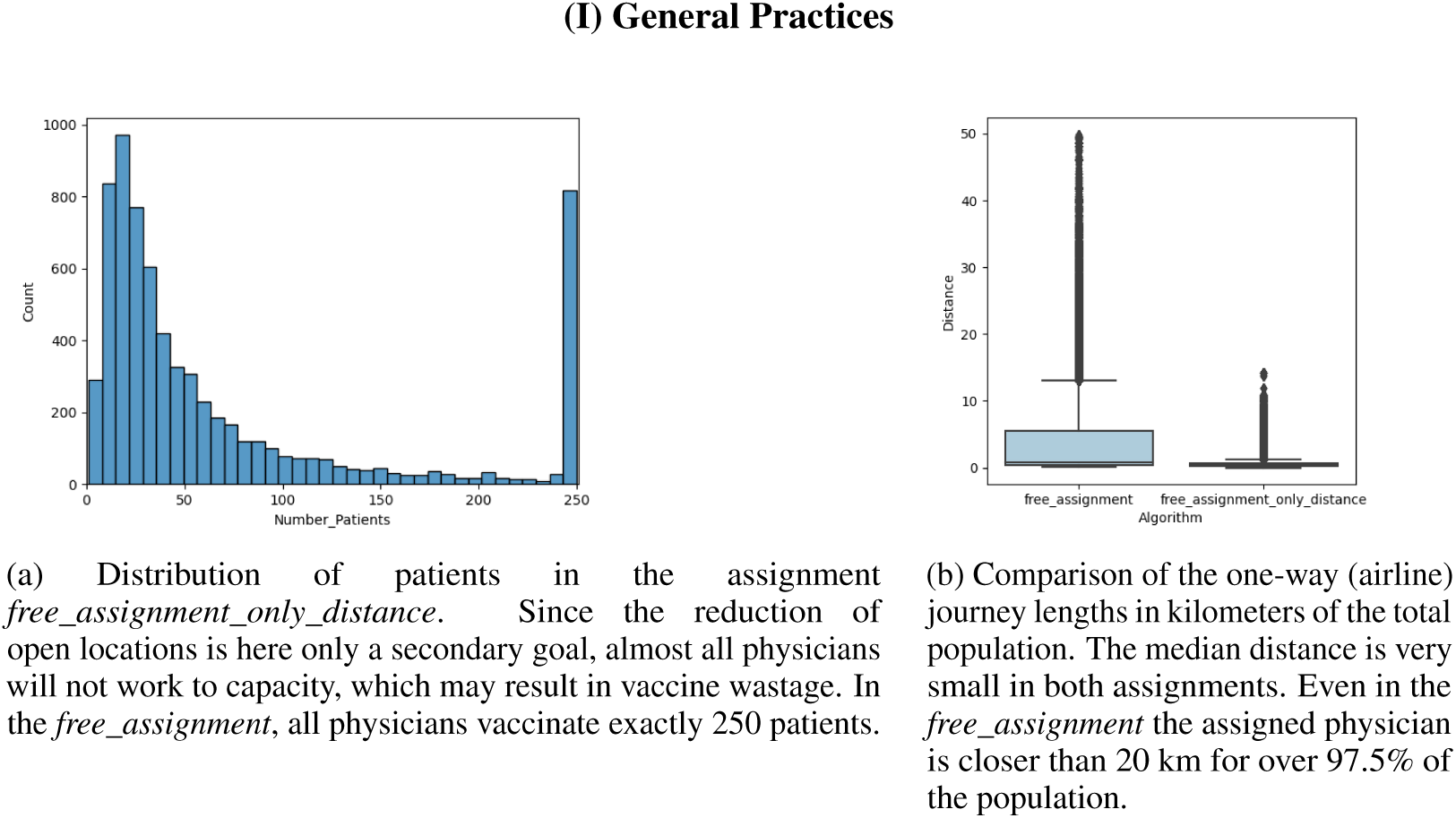

### Vaccination at Public Health Departments (II)

As previously described, each health department is responsible for a specific region and the contained municipalities. Therefore, we consider the following assignments for health departments:

i. ***closest_station_greedy***
ii. ***responsible_station***

Both assignments (i) and (ii) are fixed and only the number of necessary doctors and the distribution of distances need to be calculated.

Figure 3 shows the location of all health departments on a map. As previously, each municipality is colored according to its travel distance to the assigned health department. Note that here, for each municipality, its assigned health department is unique for both assignments. Comparing the two maps shows that the distances are quite similar in the southern and western part of Germany, but increase in the northern and eastern part. Especially the distances for the patients in the southeastern part of Mecklenburg-Vorpommern increase significantly. This is also shown in Figure 5a where we plotted the travel distances for both assignments for each federal state separately. Figure 4a shows the increase in distances aggregated for Germany.

**Figure 3:**
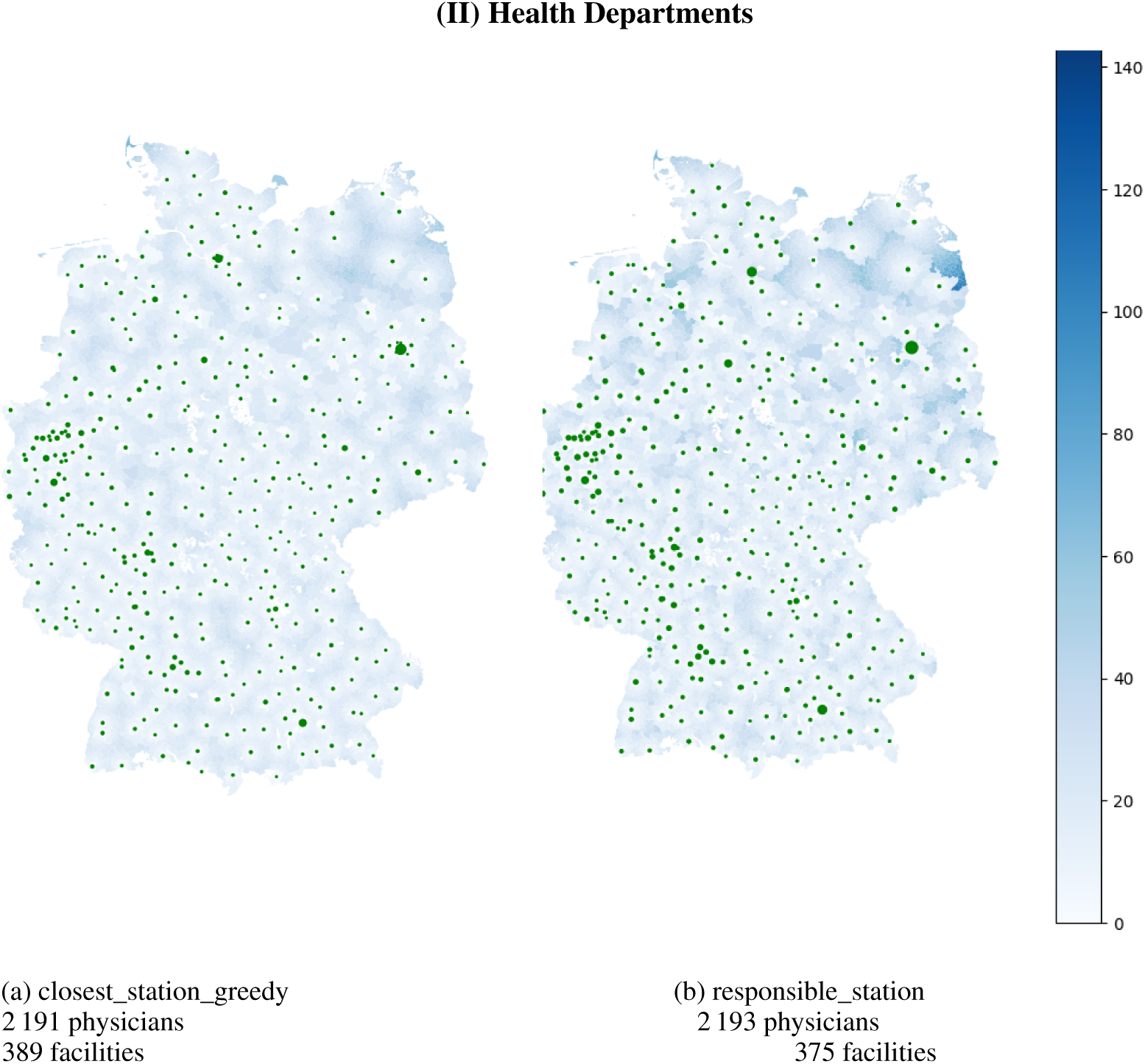
Visualization of the distance per municipality in kilometer airline for assignments (i) and (ii) in the health department scenario. Due to rounding issues, some very small municipalities are not assigned to any facility center and are kept white. In Figure 3b, some municipalities significantly increase their distance as their reference point is much closer to a health department of another county than to their responsible one. For city states, some stations are not being assigned to any municipality, since no capacities are enforced. The size of the green markers are proportional to the number of assigned patients. The differences in open facilities stems from the city states that have multiple departments, but only one is responsible for the whole city in our definition.

**Figure 4:**
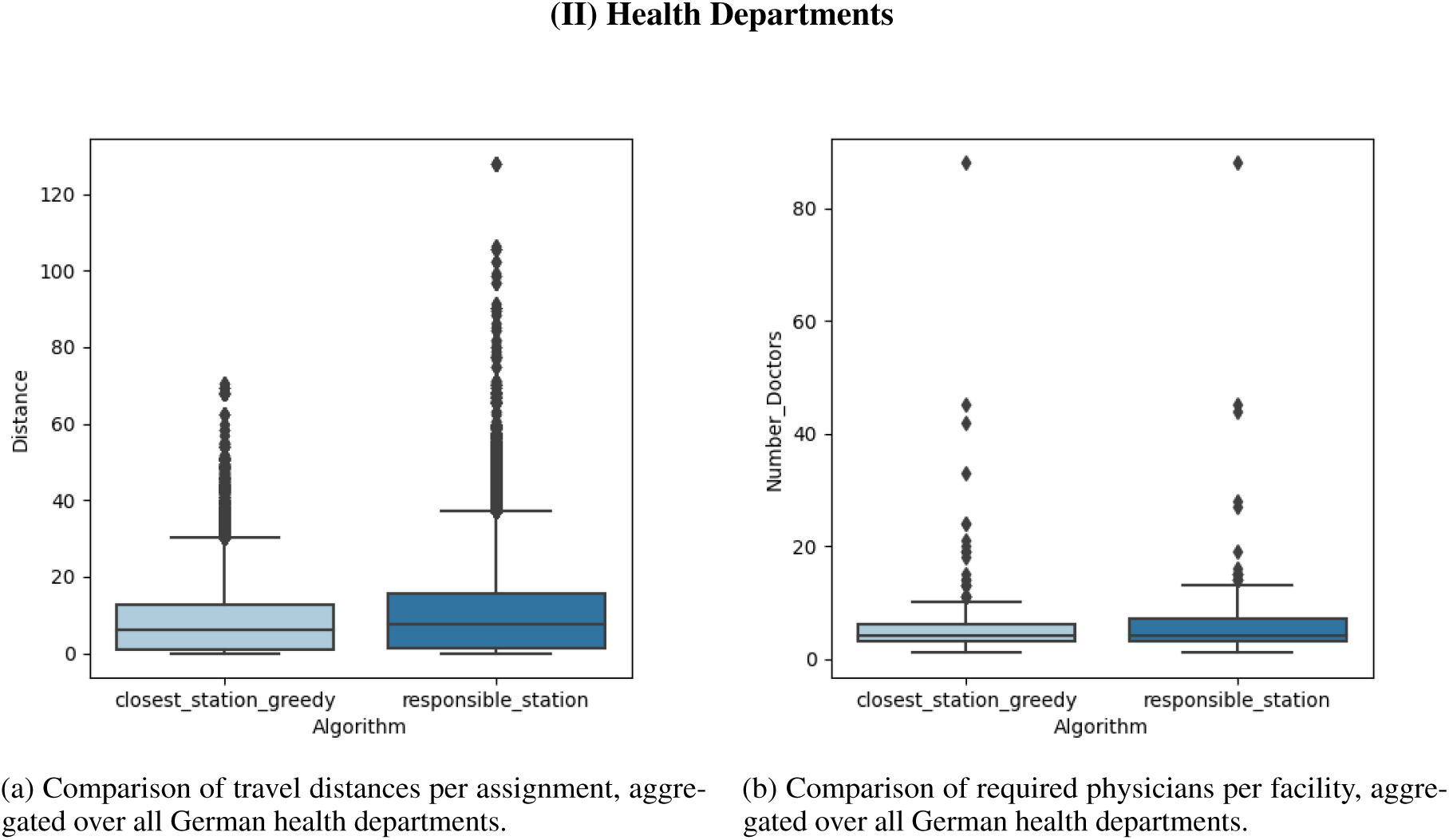
Comparison of assignments (i) and (ii) in the health department scenario aggregated over Germany. The maximal number of physicians per facility is attained in Berlin with 88 physicians, where in both scenarios the total population of Berlin is being assigned to the same health department. The maximal distance of 128 km is attained for the remote island Helgoland. However, other large distances stem also from mainland Mecklenburg-Vorpommern.

**Figure 5:**
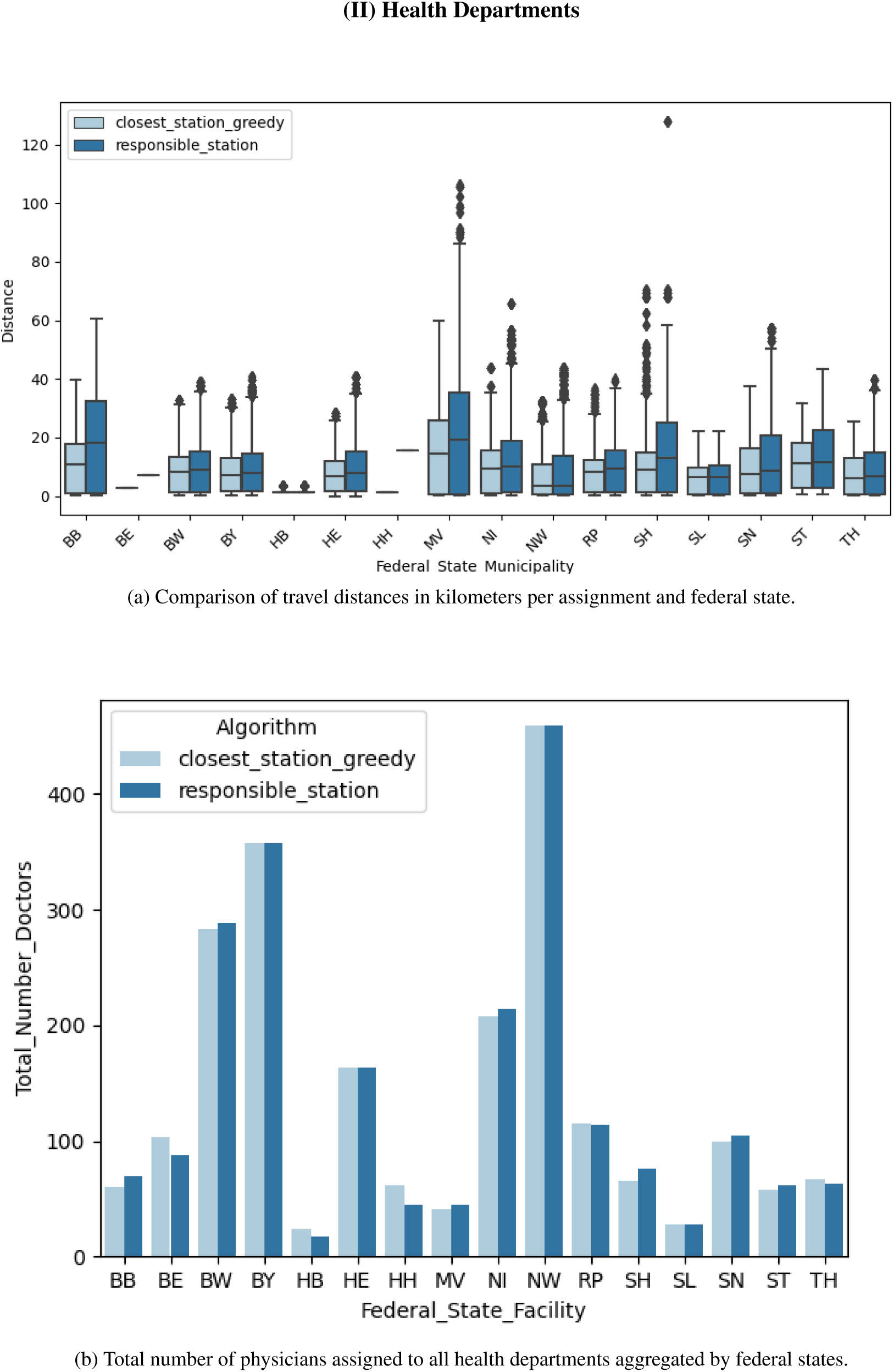
Comparison of assignments (i) and (ii) in the health department scenario. The city states naturally have the least distances. The sparsely populated Mecklenburg-Vorpommern has the most disadvantages when sticking to the administrative assignment. The number of physicians reflect the state’s population sizes.

The marker size in Figure 3 refers to the number of physicians needed at the facility to vaccine the assigned patients. Since we do not subdivide big cities with several health departments into several districts, all patients of those cities are assigned to a single health department. Therefore, in those health departments as in Berlin and Hamburg, many physicians are required whereas in practice, these physicians would be spread over several health departments. This also explains the outliers in Figure 4b where the distribution of the number of physicians per health department is shown.

The total number of physicians for the considered assignments is 2 191 and 2 193, respectively. Figures 4 and 5 shows in more detail how the physicians are distributed among Germany (4b) per facility and accumulated per federal state (5b). As expected the federal states with the most inhabitants also require the most physicians. It varies between 24, respectively 17, in Bremen and 459, respectively 459, in North Rhine-Westphalia, but the difference between the two assignments is negligible. Also the number of required physicians per health department is similar for the two assignments as shown in Figure 4b. The number of patients that are being assigned to a facility per week follows in principle the same distribution and varies from 30 to 22 200 with a median of around 1 000, i.e. 200 patients per weekday. Ignoring the outliers, 80% of all facilities would roughly face 500 to 2 000 patients per week, which translates into 2 to 4 required physicians in our setting.

### Vaccination at University Hospitals (III)

As described above, we consider three assignments for university hospitals:

i. ***closest_station_greedy***
ii. ***closest_station_same_state***
iii. ***free_assignment_only_distance***.

There are 38 university hospitals in Germany, spread over 14 of 16 federal states. Brandenburg and Bremen do not host a university hospital. Therefore, their citizens will be assigned to another university hospital within a 150 km radius.

Figure 6 shows the locations of the hospitals on a map. Additionally, each municipality is colored according to its median (w.r.t. the population) travel distance to all assigned university hospitals. Note that only in Figure 6c, a municipality can be assigned to multiple vaccination facilities. The marker sizes hint at the required number of physicians, which can be seen in more detail in Figure 7.

**Figure 6:**
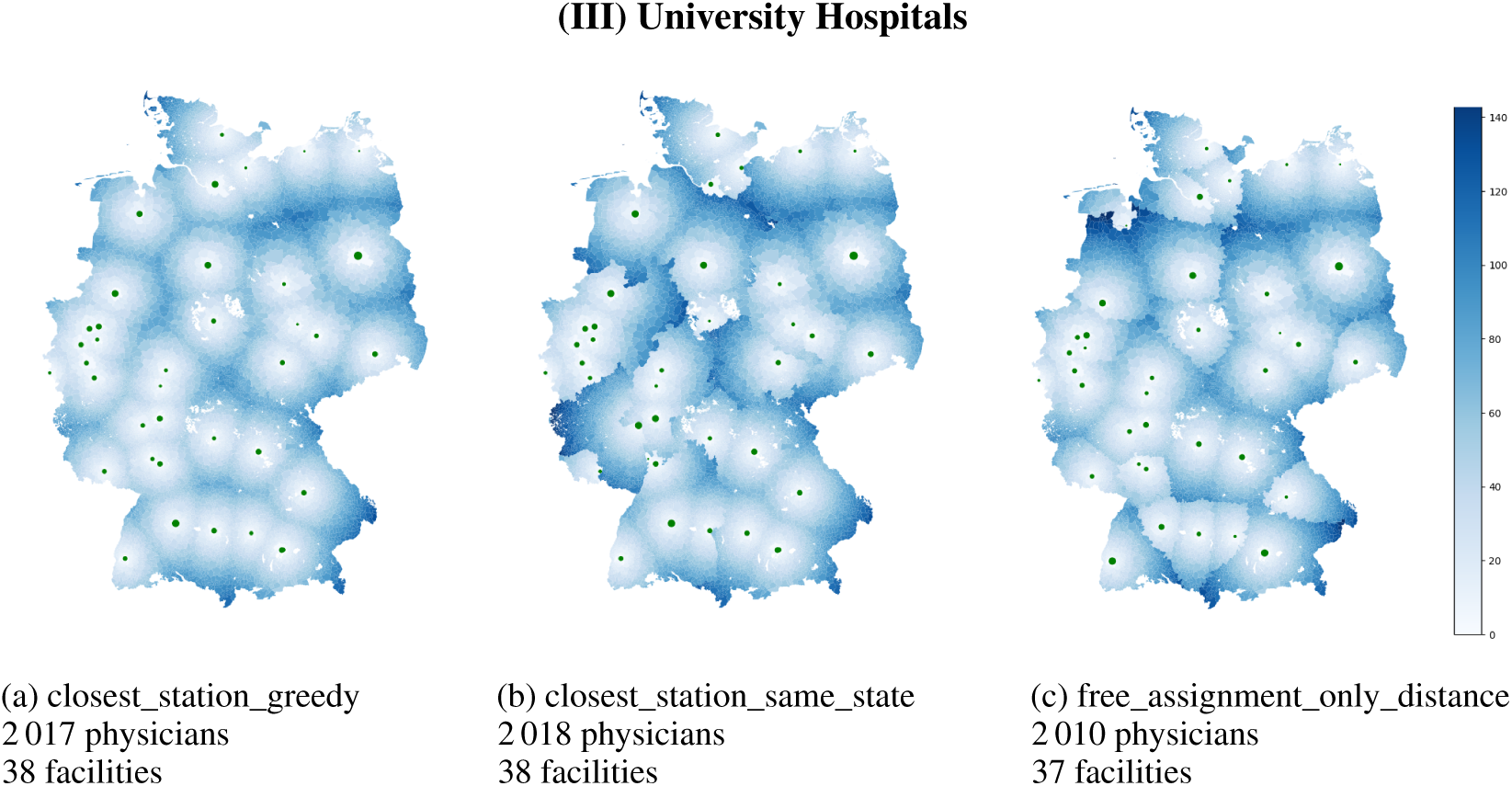
Visualization of the median distance per municipality in kilometer airline for assignments (i), (ii), and (iii) in the university hospital scenario. Due to rounding issues, some very small municipalities are not assigned to any facility center and are kept white. In Figure 6b, the borders of the German federal states are clearly visible. The marker sizes are proportional to the number of assigned patients. Note that there are two university hospitals in Munich very close by. One of them was not opened in assignment (iii) in order to save physicians.

**Figure 7:**
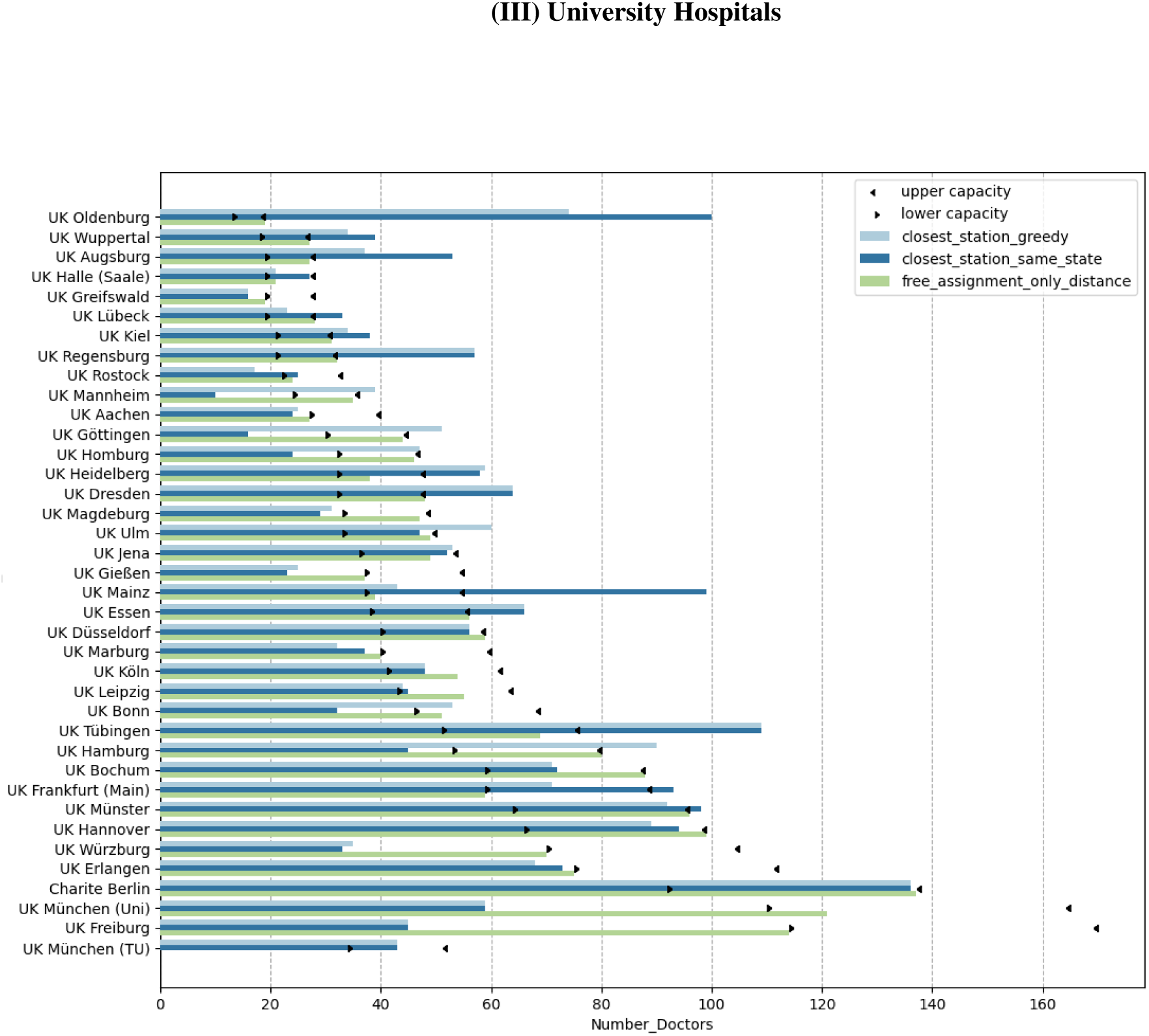
Number of vaccinating physicians assigned to each university hospital for (i) in comparison to (ii) and (iii). Only for the latter, the depicted capacity bounds had been taken into account in the model.

For assignment (i), the number of physicians per hospital vary between 16 doctors (Greifswald) and 136 doctors (Berlin). In total 2 017 doctors are required, each of whom can vaccinate up to 250 patients per week. For comparison, 2 018 doctors are needed for assignment (ii) and even 2110 doctors for assignment (iii) with capacity constraints. Note that the latter one is not necessarily solved to optimality. Our model can also compute feasible assignments with exactly 2 000 physicians, by slightly increasing the travel distances and deviate from a fixed assignment and e.g. distribute people from the same municipality to different facilities. Since the gap is rather insignificant and the distances do not alter visually, we omit this scenario for the sake of clarity.

For assignment (i) (Figure 6a), we see a circular increase in the distances around the university hospitals since everyone goes to the closest location. The map in the middle (Figure 6b) shows some substantial distance increases due to the constraint that vaccination is only allowed in the own federal state. This is especially the case for some regions in Rhineland-Palatinate, North Rhine-Westphalia and Lower Saxony. Further, the map on the right (Figure 6c) illustrates the increase of travel distance due to capacity constraints of the hospitals. The most noticeable difference can be seen around Oldenburg (Lower Saxony) in the north since the hospital capacity is much lower than the number of patients around. This is confirmed in Figure 7 where the difference between the number of physicians (which are proportional to the number of patients) in assignment (i) to (iii) are plotted. Beside the university hospital in Oldenburg, the number of patients in other hospitals such as Regensburg (Bavaria) and Tübingen (Baden-Wuerttemberg) is also reduced due to the capacity limit. In contrast to that, some hospitals such as Freiburg (Baden-Wuerttemberg) and Würzburg (Bavaria) were allocated more patients because of a lower barrier.

As university hospitals are often located in large cities, many patients reach the vaccination facility within an acceptable distance. But especially patients from rural areas have to travel a considerable distance to their facility. While Figure 6 shows the median distance of each municipality on a map, for an informed decision, it is also of interest to see the range of distances scaled by the population size. The maximum distance between a municipality center and the closest university hospital is 135 km (affecting only few people), while the median for this assignment is 30 km. Figure 8 shows boxplots of the distances for assignment (i) compared to assignment (ii) and (iii) aggregated over the total German population (Figure 8b) and grouped by states (Figure 8a). Clearly, additional constraints such as vaccination in the own federal state or capacity restrictions increase the distances travelled by the patients.

**Figure 8:**
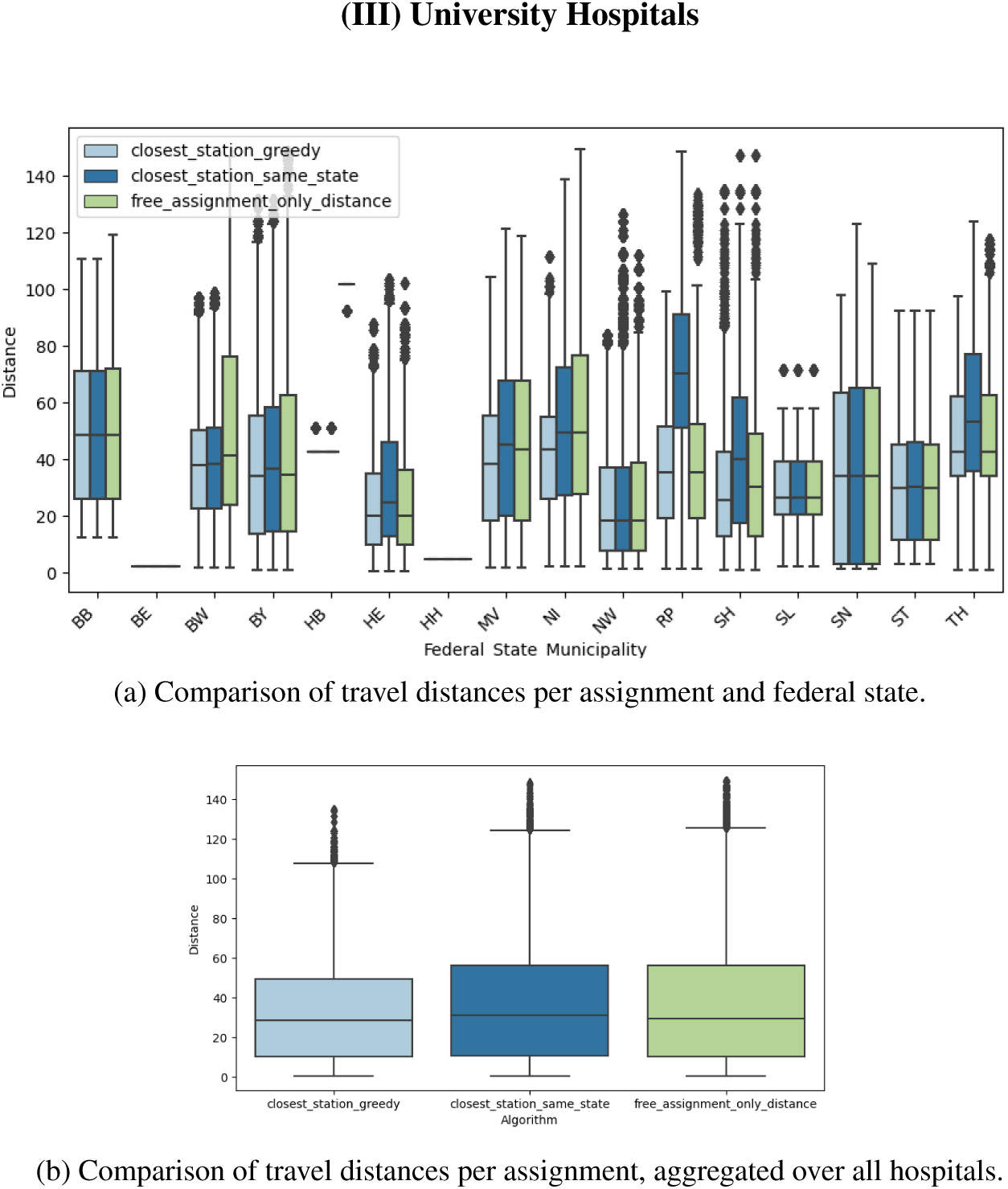
Distribution of travel distances (as linear distances in kilometers) for assignments (i), (ii), and (iii) in the university hospital scenario.

When the university hospital solution is being chosen, each facility will face a median of around 12 000 patients per week, with 80% of the facilities seeing around 6 000 to 24 000 (22 000 for assignment (i)) patients per week.

### Greenfield Planning (IV)

The last scenario we consider is an optimal choice of potential vaccination facilities such that the number of facilities and number of physicians is minimized. We allow different maximal travel distances which may only be violated if there is no other feasible assignment. More precisely we consider the assignments:

i. ***free_assignment_15km***
ii. ***free_assignment_30km***
iii. ***free_assignment_50km***
iv. ***free_assignment_75km***

Figures 9c to 9f clearly show that an even distribution of vaccination centres across Germany is optimal for minimizing distances. With an increase of the allowed travel distance, the number of required vaccination facilities decreases. On the map in Figure 3a, one can see e.g. in Mecklenburg-Vorpommern (in the north-east of Germany) that the density of health departments is rather sparse. Hence, the allowed radii of 15, 30, and 50 km can not be met for all municipalities. In Figures 9c and 9d this is reflected in darker areas mostly in the North-East. These regions are also reflected in the outliers of the boxplot in Figure 9a, where the distances of all German patients are visualized.

**Figure 9:**
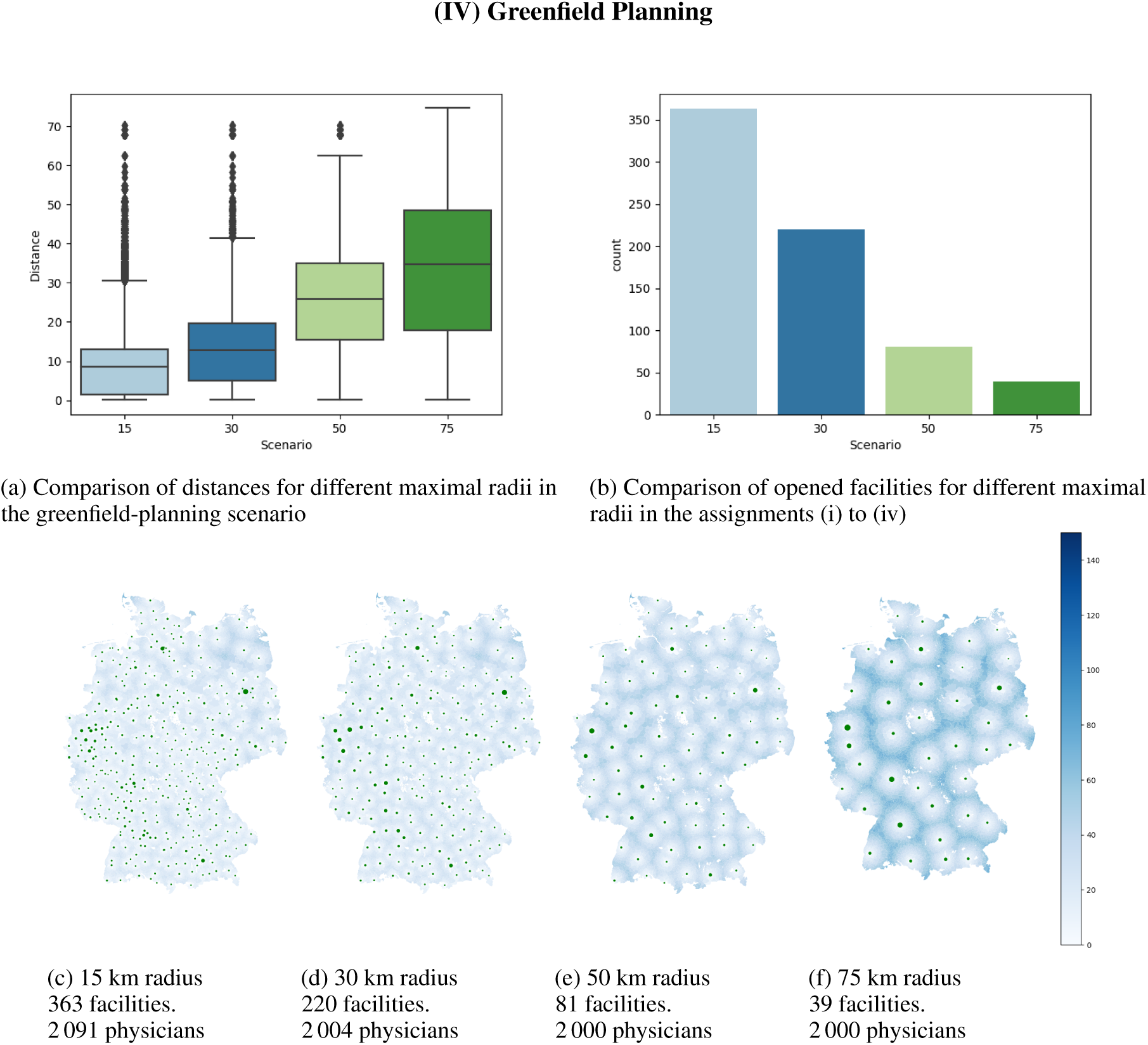
Comparison of free assignments with respect to different maximal radii. Note that municipalities whose closest facility exceeds the defined radius are being assigned to their closest facility. Therefore larger radii than allowed can occur.

For an allowed radius of 50 km, 81 facilities have to be open; for an allowed radius of 75 km, this number drops to 39. This is about the number of university hospitals, where due to a less optimized location distribution, for some patients their travel distance can be up to twice as high. Since the willingness to get vaccinated decreases with an increasing travel distance, a small radius should be chosen in practice. For an increased convenience for the patients, the required number of locations according our results rises to 220 (for 30 km airline distance) and 363 facilities for 15 km maximal allowed radius. All four assignments require up to 2 091 physicians. Due to the complexity of the model, these free assignments have not been computed up to optimality. Therefore, we cannot clearly say whether the increase in doctors for smaller radii stems from unbalanced population densities on a smaller scale or simply from unoptimal assignments.

For the sake of finding a compromise, it might however be more reasonable to not look only on maximal distances, but certain quantiles, such as the median or the 75% quantile of resulting distances, which can also be found in Figure 9a. For example, the 50 km scenario (iii) has a median distance of 25 km, i.e. 50% of all patients have a one-way journey length of 25 km or less. Looking at the 75% quantile, the distance rises to around 35 km.

## Discussion

Beyond the presented results, we have also investigated the sensitivity of the assignments with respect to several uncertain parameters. These are the composition of the population, the capacity per doctor and the total available amount of vaccine. However, all prove to have little effect in our static approach. We tested to consider only the population of persons being 65 or older. Although the proportion of seniors varies widely among German municipalities [27], our results did not show a significant variation in the structure of solutions and the performance indices of a nationwide level. However, since municipalities with a higher proportion of elderly people are typically more rural and rural areas are usually more distant from the vaccination facilities, in some states there is a shift towards an increased median travel distance and more required doctors. The variation of available doses or the variation of the capacity of one physician have in principle only linear effects in all scenarios that do not consider bounds on the vaccination facility capacity. That is, the number of required physicians roughly doubles if the number of available vaccines doubles. However, since some municipalities are very small, saturation effects on some municipalities can occur with an increased capacity. When respecting absolute capacities, an increasing number of patients obviously will change the assignment or even render it invalid when reaching the bounds. However, due to a lack of suitable data, we only considered bounds relative to the outbound patient size and by scaling these to the capacities, the solution does not alter in structure, but basically only scales the assigned patients accordingly. If data about absolute bounds is provided, it would be possible to consider this.

For the free assignments (greenfield planning), where patients are assigned to a suitable vaccination center within a prespecified radius and not all possible vaccination centers have to open, this radius of course has a huge impact on both the median travel distance as well as the location of the selected facilities. When planning the facility locations, this should be taken into account, since the COSMO Study [4] evaluated a significant drop of vaccination willingness in case the travel time (outward and return journey) exceeds 1 hour. With this in mind, our results should also motivate politicians to enable vaccination processes that are not restricted by administrative borders, since these automatically lead to avoidable peaks in travel journeys for people from border regions.

### Mathematical Background

To assign the citizens to their vaccination facility, we use two different approaches based on the model described earlier. The computation of the first and easier assignment strategy is based on a simple and fast greedy algorithm. More precisely, we determine for every region the vaccination station where the citizens of this region are vaccinated. The exact method how these stations are determined depends on which scenario we are interested in, i.e. the nearest station, the nearest station in the same state or the responsible station. Afterwards, we compute the number of people that are vaccinated in each station and determine the number of necessary physicians. In this way we solve the scenarios *closest_station_greedy, closet_station_same_state* and *responsible_station*.

The second approach is more involved and uses ideas from [12]. To compute assignment strategies for the scenarios *free_assignment* and *free_assignment_only_distance* we solve an integer program. Integer programming deals with the optimization of (linear) objective functions over a set of possible integral solutions constrained by linear equations and inequalities. For a general introduction and overview we refer to [18]. While linearity may seem like a severe restriction, together with the integrality conditions integer programming allows to model complex correlations. Integer programming is NP-complete, which means that in general there is no polynomial time algorithm, which solves all instances to optimality, unless P=NP. Nevertheless, exponential time algorithms are available. Thus, to solve the integer program corresponding to our model we use Gurobi Optimizer, cf. [9].

- As described above we use different objects for our model. To keep the overview we summarize all these objects here.
- **The set J of regions:** A set of areas where the citizens to be vaccinated live (e.g., municipalities).
- **The set I of vaccination stations:** A set of possible vaccination centers (e.g., general practices, health departments, university hospitals).
- **The neighborhood N**(**j**) **of the region j** ∈ **J:** Possible vaccination centers that are responsible for the region *j* (e.g., determined by distance or predefined assignment).
- **The neighborhood N**(**i**) **of the station i** ∈ **I:** Regions from which citizens can be vaccinated in vaccination center *i* (e.g., determined by distance or a predefined assignment).
- **Number of citizens d**_**j**_ **to be vaccinated in region j (**e.g., 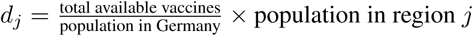)
- **Maximal capacity of vaccinations cap**_**i**_ **in station i I** (e.g., can be set to infinity or another predefined value)
- **Lower Bound of vaccinations lb**_**i**_ **in station i** ∈ **I if station** *i* **is opened as a vaccination center** (e.g., can be set to 0 or another predefined value)
- **Maximal number of vaccinations b that can be carried out by a physician** (e.g., can be set to 250)

The remarks in the brackets are suggestions for these values and we mainly deal with these stated possibilities. Observe that these values always refer to the time frame that is fixed, in our case these values correspond to a 5-day week with 8 working hours each. However, if the appropriate data is available, these values can be replaced at will. This shows that the model is very universal and adaptable for different scenarios.

In the integer program we use three different variables. In the following we describe their purpose:

- **x**_**i**_ ∈{0, 1} for *i* ∈ *I*: For a vaccination center *i* ∈ *I* the variable *x*_*i*_ states in a solution whether the vaccination center *i* is opened (*x*_*i*_ = 1) or closed (*x*_*i*_ = 0).
- **y**_**i**_ ∈ N for *i* ∈*I*: For a vaccination center *i* ∈*I* the variable *y*_*i*_ states in a solution the number of physicians needed in vaccination center *i*.
- **z**_**i**,**j**_ ∈N for *i* ∈*I* and *j* ∈*J*: For a vaccination center *i* ∈*I* and a region *j* ∈*J* the variable *z*_*i,j*_ states in a solution how many citizens from region *j* are vaccinated in vaccination center *i*.

Thus, in a solution (*x, y, z*) variable *x* yields an answer to question 1, variable *y* gives an answer to question 2 and variable *z* implies an answer to question 3. We are now ready to present the integer program, where we abbreviate the objective function with a placeholder function *f* (*x, y, z*) = *a*^*T*^ *x* + *b*^*T*^ *y* + *c*^*T*^ *z*:

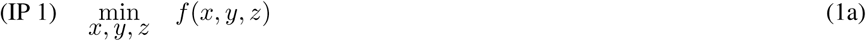

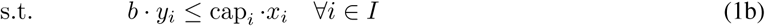

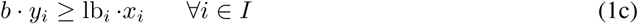

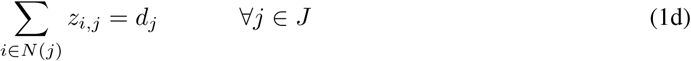

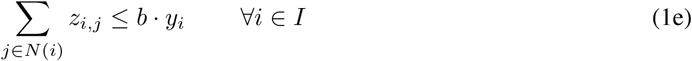

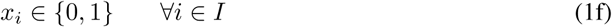

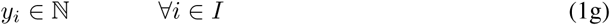

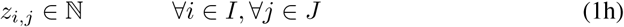

In (IP 1) constraint (1b) ensures that if a vaccination center is open (i.e., *x*_*i*_ = 1), then there are at most as many possible vaccinations by the physicians as the capacity cap*i* of the vaccination center *i* allows. In particular, this bounds the number of physicians in each vaccination center. Constraint (1c) makes sure that if a vaccination center *i ∈ I* is open, then at least lb_*i*_ vaccinations are carried out in *i*. In constraint (1d), we guarantee that, in each region *j*, exactly *d*_*j*_ citizens are being vaccinated. Note that, in some scenarios, these citizens are vaccinated in different facilities. Constraint (1e) ensures that all vaccinations in vaccination center *i* can be performed by the physicians assigned to *i*. The model is a variant of a *facility location problem*. For a general overview on facility location we refer to [23, 8]. Another facility location model used to select pharmacies for Covid19-testing is given in [17]. For a more recent survey on health care related facility location problems we refer to [1].

The model above is quite universal, different objective functions allow to focus on various goals. We have implemented various objective functions and give a short overview in the following:

- **Minimizing the number of open vaccination stations:**

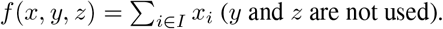
- **Minimizing the number of needed physicians:**

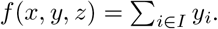
- **Minimize the sum of the travel distances of the patients**

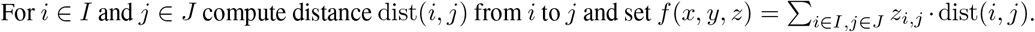

It is important to note that we can optimize not only a single one of these objective functions, but also multiple of them in any lexicographical combination. If, for example, our given combination is (a) and (b), this means that when optimizing objective (a) we only consider optimal solutions of objective (b) as possible solutions. Therefore, we obtain different assignment strategies for the doctors and information which patient should be vaccinated at which vaccination center in our scenarios.

### Outlook

The situation of planning a population-wide vaccination campaign against a deadly virus several months in advance of the approval of any vaccine in the middle of a pandemic is unique in history. It is therefore not surprising that the assumptions and knowledge of many parameters regarding the availability and shelf life of vaccine doses and the technology required are constantly changing. The present study can thus only reflect the current state of discussion. It is also only suitable to decide on the question of the type of vaccination locations and gives a rough allocation of citizens to places.

The tactical planning gives rise to new questions. Instead of optimizing the conflicting objectives lexicographically, it could be beneficial to analyse them in a multi-objective optimization approach, in order to highlight good compromises that, e.g., increase the distances only for very few people while balancing the required number of doctors at the individual facilities. Another aspect that could be taken into account is the heterogeneous willingness in the population to be vaccinated, which on the one hand possibly depends on the local occurrence of infection, but on the other hand also on the distance to the vaccination site.

Evaluating the vaccination assignment as a time-dynamic organizational problem may also include varying vaccine availability over time, changing demand in the population and a potential need of revaccinating patients after a certain number of weeks or months.

The operational planning of such a vaccination campaign must then be carried out at the state level. This allows also for taking regional characteristics and actual road distances into account. It should also be stated that a more refined location planning would also compensate for the flaw of very different definitions of a municipality throughout Germany in terms of area and population size.

## Data Availability

available upon request

## Author’s contributions

NL and JS analyzed the data, created the plots and had major parts in defining the research questions. ES, MS and SK designed the mathematical model. SJ, MS and ES performed the computations. NL, JS, SJ, MS and ES contributed text to the paper. SS contributed data and details about the planning state and has made substantial contributions to the design of the research questions. All authors read and approved the final manuscript.

## Acknowledgements

This work was partially supported by the German Federal Ministry of Education and Research within the project “HealthFaCT-Cor - Health: Facility Location, Covering and Transport - Corona”.

## References

[1] Amir Ahmadi-Javid, Pardis Seyedi, and Siddhartha S Syam. A survey of healthcare facility location. Computers & Operations Research, 79:223–263, 2017.

[2] Nationale impfstrategie covid-19. Accessed: 2020-11-12.

[3] Xin Chen, Menglong Li, David Simchi-Levi, and Tiancheng Zhao. Allocation of COVID-19 vaccines under limited supply. aug 2020.

[4] Cosmo — covid-19 snapshot monitoring, 2020.

[5] Dhl white paper - resilienz in pandemien. Accessed: 2020-11-12.

[6] EU strategy for covid-19 vaccines. Accessed: 2020-11-12.

[7] Preparedness for covid-19 vaccination strategies and vaccine deployment, oct 2020. Accessed: 2020-11-12.

[8] Reza Zanjirani Farahani, Maryam SteadieSeifi, and Nasrin Asgari. Multiple criteria facility location problems: A survey. Applied mathematical modelling, 34(7):1689–1709, 2010.

[9] LLC Gurobi Optimization. Gurobi optimizer reference manual, 2020.

[10] Corona-forschung am HCHE. Accessed: 2020-11-12.

[11] Indikatoren und karten zur raumund stadtentwicklung. Accessed: 2020-11-12.

[12] Sven O. Krumke, Eva Schmidt, and Manuel Streicher. Robust multicovers with budgeted uncertainty. European Journal of Operational Research, 274(3):845–857, may 2019.

[13] Infoportal zukunft.land. Accessed: 2020-11-12.

[14] Jeffrey V. Lazarus, Scott C. Ratzan, Adam Palayew, Lawrence O. Gostin, Heidi J. Larson, Kenneth Rabin, Spencer Kimball, and Ayman El-Mohandes. A global survey of potential acceptance of a COVID-19 vaccine. Nature Medicine, oct 2020.

[15] Laura Matrajt, Julie Eaton, Tiffany Leung, and Elizabeth R Brown. Vaccine optimization for COVID-19, who to vaccinate first? aug 2020.

[16] Coronavirus: Der große impfplan für deutschland. Accessed: 2020-11-12.

[17] Simon Risanger, Bismark Singh, David Morton, and Lauren Ancel Meyers. Selecting pharmacies for COVID-19 testing to ensure access. sep 2020.

[18] Alexander Schrijver. Theory of linear and integer programming. John Wiley & Sons, New York, NY, 1998.

[19] Seaborn documentation of boxplots. Accessed: 2020-11-12.

[20] Wie hält man 1,3 milliarden dosen bei minus 70 grad? Accessed: 2020-11-12.

[21] Statistisches bundesamt. Accessed: 2020-11-12.

[22] Graham Upton and Ian Cook. Understanding statistics. Oxford University Press, Oxford, UK, 1996.

[23] Vedat Verter. Uncapacitated and capacitated facility location problems. In H. A. Eiselt and Vladimir Marianov, editors, International Series in Operations Research & Management Science, pages 25–37. Springer US, Boston, MA, 2011.

[24] Roadmap ffor prioritizing population groups for vaccines against covid-19. Accessed: 2020-11-12.

[25] Who sage values framework for the allocation and prioritization of covid-19 vaccination. Accessed: 2020-11-12.

[26] Daten der arztsuchen, 2013. Accessed: 2020-11-12.

[27] Zensus2011, 2011.

